# Sources of Firearms Used by Children in K-12 School Shootings, 1999-2021

**DOI:** 10.1101/2023.12.16.23300073

**Authors:** Vivian V. Studdert, Matthew Miller

## Abstract

**Background:** Shootings in US schools have become common and deadly events. Little is known about the sources of firearms used in these shootings.

**Methods:** We used the K-12 School Shooting Database (K-12 SSDB) to identify a sample of shootings on school campuses between 1999 and 2021 that were perpetrated by children (<18 years) and resulted in at least one death or injury. We searched news stories and public records to gather information on the sources of the firearms used in these shootings. Additional descriptive information on the shootings, shooters, and victims came from the K-12 SSDB.

**Results:** A total of 217 school shootings met our eligibility criteria, and we recovered information on the source of the firearms used in 65 (30%) of them. Most of the shootings occurred inside (68%) and involved one shooter (97%) who typically used a handgun (80%). A total of 71% of the shootings involved assault or homicide, 26% were accidental, and 3% involved an assault or homicide that ended in the shooter’s suicide. In 60% of the shootings, shooters obtained the firearm(s) from their home. The next most common sources were another person’s home (17%) or the street (17%). Among shootings for which the firearms were obtained from the shooter’s home, 77% belonged to a parent or guardian and 18% belonged to another relative.

**Conclusions:** Firearms used by child perpetrators of K-12 school shootings come predominantly from the shooter’s home, where they usually belong to the perpetrator’s parents. Stronger adherence to guidelines for safe firearm storage has the potential to reduce risks of these tragedies.

## Introduction

Firearm violence—and apprehension and fear of firearm violence—have become pervasive features of K-12 education in the United States. Dozens of shootings occur on school campuses each year.^1^ Over the past 20 years, these incidents, most of which are perpetrated by adolescents,^2^ have killed or wounded thousands of students and staff.^1^ Countless more people survive without physical wounds but experience lasting trauma.^3^

Contrary to the picture presented by school shootings that receive widespread media coverage, most school shootings are not mass casualty events involving assault-style rifles. Rather, they typically involve handguns and interpersonal disputes, mirroring broader patterns of lethal assaults in the US; moreover, many are perpetrated by people who are not students of the school, occur outside of school buildings (e.g., yard, stadium) during non-school hours, and are gang-related.^2,4,5,,6^

Little is known about the sources of firearms used in school shootings. We identified only one empirical study in the scientific literature, a recent analysis by Klein et al^4^, that describes where perpetrators of school shootings obtained the guns they used. The provenance of firearms used by shooters who are younger than 18 years is of particular interest because it is illegal in all states for children to purchase or possess handguns, and there are restrictions under federal and state law on children’s access to long guns.^7^

Using a different database than the one used by Klein et al, supplemented by manual searches of public records and media reports, we identified the sources of firearms used in a sample of school shootings perpetrated by children between 1999 and 2021.

## Methods

We used the K-12 School Shooting Database (“K-12 SSDB”)^1^ to identify shootings that occurred between April 20, 1999 and December 31, 2021. This database compiles information on school shootings since 1970 using open source information (government reports, media outlets, other tracking databases). Inclusion criteria are expansive, covering instances when “a gun is fired, brandished…, or bullet hits school property, regardless of the number of victims, time, day, or reason”.^1^

The time period for our study sample began on the date of the Columbine High School shooting and extended through 2021. Despite their availability in the K-12 SSDB, we did not include more recent shootings because our preliminary investigations showed that reports related to subsequent court proceedings were often an important contributor of information, and these were generally unavailable until several years after the shootings occurred. Shootings in our study time period that met the following criteria entered our study sample: occurred on a K-12 school campus, resulted in at least one injury or death, shooter(s) aged <18 years, use of a firearm with explosive propellant (as opposed to BB guns, pellet guns, etc.), and the shooter was apprehended or identified.

The source of firearms used in the shootings, our variable of interest, is not recorded in the K-12 SSDB. We searched for it in public records and media reports. Our primary source was the Lexis+ news database, which we searched using the following Boolean terminology: *[“[school name]” w/5 “[school type – e*.*g*., *middle school]”] AND “[city name]” AND (shooting OR shot OR gun OR firearm)*. We also searched Google using similar terms, and cross-referenced the Washington Post school shootings database^8^ (a tracker with much more selective inclusion criteria than the K-12 SSDB) which included at least some source information on approximately 40% of the shootings in our sample.

All information on sources recovered in these searches was transcribed. Next, based on a review of source information associated with 25 randomly selected shootings, we created a preliminary taxonomy of sources. The taxonomy was further refined and finalized during review of 25 more shootings. One reviewer (VS) then coded the rest of the sources according to the final taxonomy. The coding was completed in summer 2023.

Other descriptive information on the shootings, shooters, and firearms (e.g., demographics, number of shooters, location, type of firearm) came chiefly from the K-12 SSDB.

## Results

A total of 217 school shootings met our eligibility criteria, and we were able to recover information on the source of the firearms used for 65 (30%) of them (**Figure 1**). All results reported hereafter relate to those 65 shootings, which involved 67 shooters and 174 victims (46 with fatal injuries, 128 with non-fatal injuries).

**Figure 1.**
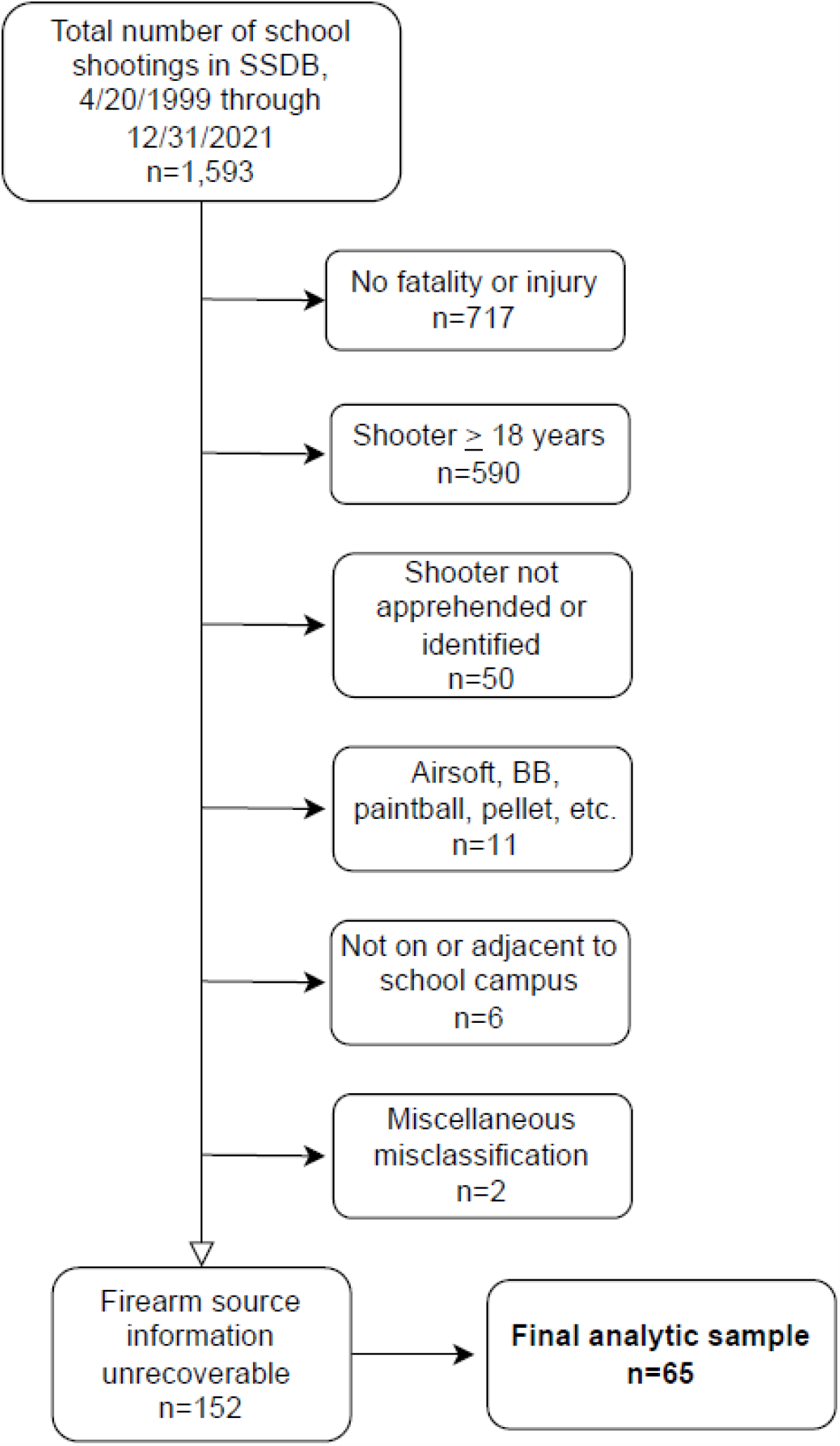
Derivation of study sample *.

**\* For shootings with multiple shooters, the “shooter ≥ 18 years” exclusion category applied if any of the shooters were aged 18 years or older.**

**Table 1** provides descriptive characteristics of the shootings, shooters, and victims. Most of the shootings occurred inside (68%) and involved one shooter (97%), who typically used a handgun (80%). A total of 71% of the shootings involved assault or homicide, 26% were accidental, and 3% involved an assault or homicide that ended in the shooter’s suicide.

**Table 1.**
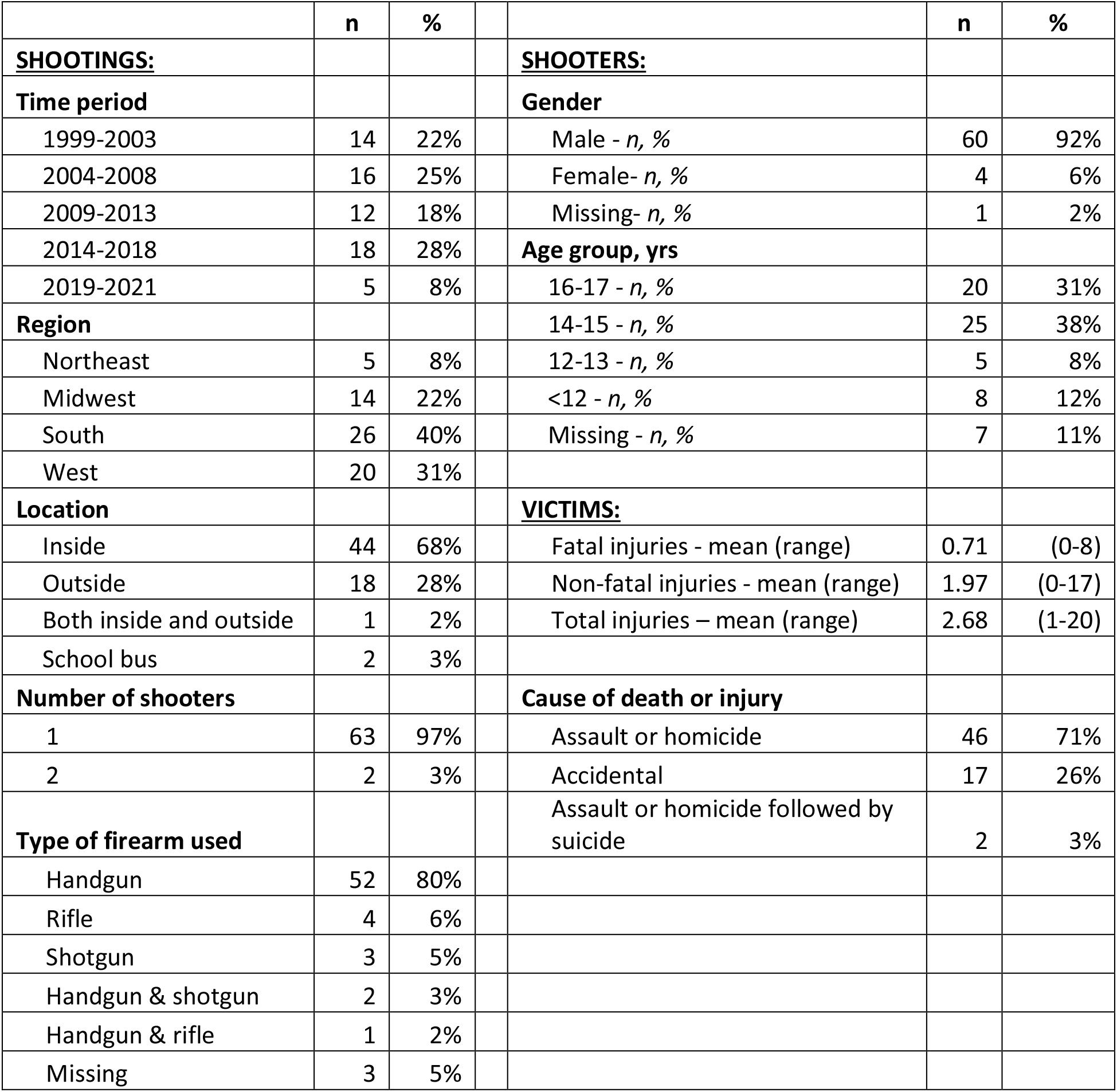
Characteristics of shootings and shooters in study sample.

A total of 92% of the shooters were male and nearly a quarter of those whose age was reported were aged 13 years or younger.

In 60% of the shootings, shooters obtained the firearm(s) from their home (**Table 2**). The next most common sources were another person’s home (17%), such as a friend’s or relative’s, and the street (17%), where shooters obtained firearms through various methods, including illegal purchases, theft, and finds.

**Table 2.** Sources of firearm used in school shooting.

| **Physical location** | *n* | % |  | **Owner of firearm shooter**<br>**obtained from shooter's home** | *n* | % |
| --- | --- | --- | --- | --- | --- | --- |
| Shooter's home | 39 | 60% |  | Parent | 30 | 77% |
| Other home | 11 | 17% |  | Other relative | 7 | 18% |
| Street | 11 | 17% |  | Friend or acquaintance | 1 | 3% |
| Store or pawn shop | 3 | 5% |  | Unknown | 1 | 3% |
| Vehicle | 1 | 2% |  |  |  |  |

Among shootings for which the firearms were obtained from the shooter’s home, 77% belonged to a parent or guardian and 18% belonged to another relative. Thus, overall, shooters’ guns were acquired from home and belonged to a parent, guardian, or relative of the shooter in 57% of all shootings in our sample.

## Discussion

This study found that firearms used by child perpetrators of K-12 school shootings came predominantly from the shooter’s home, where they usually belonged to the shooter’s parents.

Our study extends what is, to our knowledge, the only other published study of sources of weapons used in school shootings. We relied on a different database than Klein et al.^5^ In some respects our sample eligibility criteria were broader—for example, we did not exclude *a priori* shootings that resulted in injuries from firearm accidents or suicide attempts, and our study window extended through 2021. In other respects, our eligibility criteria were narrower—for example, we excluded 19-year-olds and shootings that did not result in injury or death. Nonetheless, despite these differences, our finding that 57% of shooters used firearms that were obtained in their home and owned by a parent, guardian or relative was similar to Klein at al’s finding that 54% of shooters in their sample had acquired their guns from family members or relatives.

Our study has several limitations. Information on the sources of weapons could not be recovered in 75% of the shootings that met our eligibility criteria. (Klein et al could not recover sources in 51% of shootings in their sample.^5^) When available, this information often came from reports of investigations or court cases months or years after the event, but these reports tended to be sparing with details, possibly out of concern for the shooter’s status as a minor. In addition, even when information on provenance was recoverable, it did not routinely provide important details, such as the extent to which the shooters’ parents had authorized or enabled the child’s access, or how shooters who obtained weapons on the street did so.

A recent national survey estimated that guns are stored unlocked in nearly half of all households that have both firearms and children.^9^ Approximately 25 states have enacted child-access prevention (CAP) or safe storage laws to deter owners from leaving firearms accessible to minors.^10^ Given the predominance of weapons in the home as a source of guns for children who perpetrate school shootings, adherence to guidelines for safe firearm storage, coupled with wider adoption, stronger enforcement, and better promotion of the rationale undergirding CAP and safe storage laws, have potential to reduce the risks of these senseless tragedies.

## Data Availability

All data produced in the present study are available upon reasonable request to the authors.

https://k12ssdb.org/

## Acknowledgements

The authors thank David Riedman for providing an extract of the K-12 School Shooting Database.

## References

1. Riedman D. K-12 School Shooting Database. https://k12ssdb.org/

2. Freilich JD, Chermak SM, Connell NM, Klein BR, Greene-Colozzi EA. Using open-source data to better understand and respond to American school shootings: introducing and exploring the American School Shooting Study (TASSS). J Sch Violence. 2022;21(2):93–118.

3. Cabral M, Kim B, Rossin-Slater M, Schnell M, Schwandt H. Trauma at school: The impacts of shootings on students’ human capital and economic outcomes. Working paper 28311, December 2020 https://www.nber.org/papers/w28311 (accessed December 2023)

4. Gammell SP, Connell NM, Huskey MG. A descriptive analysis of the characteristics of school shootings across five decades. Am J Crim Justice. 2022;47(5):818–835.

5. Klein BR, Freilich JD, Chermak SM. K-12 school shootings in context: New findings from the American School Shooting Study (TASSS). Rockefeller Institute of Government, August 2023.

6. Freilich JD, Connell NM, Klein BR, Greene-Colozzi EA. Overview of the American School Shooting Study (TASSS). Rockefeller Institute of Government; August 2022.

7. Giffords Law Center. Minimum age to purchase & possess. https://giffords.org/lawcenter/gun-laws/policy-areas/who-can-have-a-gun/minimum-age/ (accessed December 2023).

8. Washington Post. Schools shootings data. https://github.com/washingtonpost/data-school-shootings?itid=lk_inline_enhanced-template (accessed December 2023).

9. Miller M, Azrael D. Firearm storage in US households with children: Findings from the 2021 National Firearm Survey. JAMA Network Open 2022;5(2):e2148823.

10. Giffords Law Center. Child access prevention and safe storage. https://giffords.org/lawcenter/gun-laws/policy-areas/child-consumer-safety/child-access-prevention-and-safe-storage/ (accessed December 2023).

